# Patient Attitudes Toward Artificial Intelligence in Cancer Care: A Scoping Review

**DOI:** 10.1101/2025.03.15.25324029

**Authors:** Daniel Hilbers, Navid Nekain, Alan T. Bates, John-Jose Nunez

## Abstract

**PURPOSE:** To synthesize existing literature on patient attitudes toward AI in cancer care and identify knowledge gaps that can inform future research and clinical implementation.

**DESIGN:** A scoping review was conducted following PRISMA-ScR guidelines. MEDLINE, EMBASE, PsycINFO, and CINAHL were searched for peer-reviewed primary research studies published until February 1, 2025. The Population-Concept-Context framework guided study selection, focusing on adult patients with cancer and their attitudes toward AI. Studies with quantitative or qualitative data were included. Two independent reviewers screened studies, with a third resolving disagreements. Data were synthesized into tabular and narrative summaries.

**RESULTS:** Our search yielded 1,240 citations, of which 19 studies met the inclusion criteria, representing 2,114 patients with cancer across 15 countries. Most studies used quantitative methods (n=9) such as questionnaires or surveys. The most studied cancers were prostate, melanoma, breast, and colorectal cancer. While patients with cancer generally supported AI when used as a physician-guided tool, concerns about depersonalization, treatment bias, and data security highlighted challenges in implementation. Trust in AI was shaped by physician endorsement and patient familiarity, with greater trust when AI was physician-guided. Geographic differences were observed, with greater AI acceptance in Asia, while skepticism was more prevalent in North America and Europe. Additionally, patients with metastatic cancer were underrepresented, limiting insights into AI perceptions in this population.

**CONCLUSION:** This scoping review provides the first synthesis of patient attitudes toward AI across all cancer types and highlights concerns unique to patients with cancer. Clinicians can use these findings to enhance patient acceptance of AI by positioning it as a physician-guided tool and ensuring its integration aligns with patient values and expectations.

## Introduction

Artificial intelligence (AI) is improving cancer care by advancing detection, diagnosis, and treatment planning.^1–3^ These technologies are being applied to improve enhance diagnostic accuracy, predict survival outcomes, and personalize treatment strategies.^4–11^ However, AI’s impact depends not only on its technical capabilities but also on patient acceptance and trust. Understanding these perspectives is essential to ensuring a successful patient-centered approach to AI implementation.^14^

While general attitudes of all patients toward AI have been studied, research specifically examining the perspectives of patients with cancer remains limited.^15^ The chronic and severe nature of cancer creates unique psychosocial challenges for patients, which may shape their attitudes toward AI in ways distinct from other medical contexts.^12,13^ The limited existing studies primarily focus on individual cancer types, such as breast and skin cancer^16–19^, leaving gaps in understanding how AI is perceived across diverse cancer care contexts. A more comprehensive evaluation is needed to capture the full spectrum of patients’ attitudes toward AI in cancer care.

This scoping review maps the existing literature, identifies knowledge gaps, and highlights opportunities for future research regarding patients’ attitudes toward AI in cancer care. Given its broad and exploratory scope, this review provides a foundation for guiding the patient-centered development and implementation of AI in cancer care.^20–22^

## Methods

This scoping review followed a six-stage methodological framework and adhered to the Preferred Reporting Items for Systematic Reviews and Meta-Analyses guideline (PRISMA-ScR).^21,23^ The review was registered with Open Science Framework (https://osf.io/8zph9) and was conducted according to the published protocol.^24^

## Search Strategy

To formulate the research question, we applied a conceptual framework for evaluating patient attitudes toward AI in healthcare.^25^ Following Joanna Briggs Institute’s (JBI) recommended three step search strategy, we conducted an initial limited search of MEDLINE and EMBASE to identify key words and relevant index terms.^20–23,26^ These keywords and index terms were used to develop a final search strategy and to conduct a literature search across the identified databases for this review. The MEDLINE, EMBASE, PsycINFO, and CINAHL databases were searched for English-language primary qualitative and quantitative studies published in peer-reviewed journals until February 1, 2025. The reference lists of literature included in this scoping review were searched for additional relevant studies. The final search strategy, found in eMethods in the Supplement, was developed with support from subject librarians at the University of British Columbia.

## Inclusion and exclusion criteria

The eligibility criteria were determined using the Population-Concept-Context framework.^26^

### Population

This review included adult patients with cancer with all forms and stages of cancer. For mixed populations of patients with and without cancer, we included studies if the patients with cancer population was the majority, or if the population is composed of those with cancer and those with non-malignant tumours receiving specialized care. Also, we included mixed populations of patients with cancer and physicians or caregivers, only if patient attitudes were reported independently. For a study to be included, AI must have been involved, and we defined AI as a computer system modelling intelligent behaviour with minimal human intervention.^27^

### Concept

Attitudes on the use of AI in cancer care. We defined attitudes as input received from patients that can broadly be understood as thoughts, feelings, emotions, perspectives, attitudes, opinions, sentiments, beliefs, and experiences.

### Context

We included English-language primary qualitative and quantitative studies published in peer-reviewed journals until February 1, 2025 and included all geographies and clinical settings.

## Data screening, Extraction and Synthesis

The searches identified 1,641 sources, of which 401 were duplicates. This left 1,240 potential sources that were reviewed for inclusion by two independent reviewers, with discrepancies resolved by a third reviewer when necessary. Nineteen studies met the inclusion criteria. The process of study selection is detailed in Figure 1. The following data items were collected: journal name, study design, sample size, methodology, key findings, type of AI used, cancer type, and patient attitudes toward AI. Attitude themes included preferences for physician-only care, AI-only care, or physician-guided AI, as well as patient outlook, trust, satisfaction, and fear. The data was charted in Microsoft Excel and the studies were grouped by type of attitude. Table 1 provides a summary of the included studies, while details about methodology, frequency counts and key findings are provided in narrative form in eTable in the Supplement.

**Table 1.** Summary of 19 Studies Included

| Source | Study Type | AI Type | Sample Size <sup>a</sup> | Cancer Type(s) | Attitudes towards AI use in cancer care |  |  |  |  |
| --- | --- | --- | --- | --- | --- | --- | --- | --- | --- |
|  |  |  |  |  | AI vs Physician <sup>b</sup> | Patient Outlook <sup>c</sup> | Trust <sup>c</sup> | Satisfaction <sup>c</sup> | Fear <sup>c</sup> |
| Au et al, 2023 <sup>28</sup> | Case study | Chatbot | 1 | Choroidal metastasis | NA | NA | NA | + | NA |
| Fransen et al, 2025 <sup>40</sup> | Cross-sectional study | Predictive | 206 | Prostate Cancer | Combined | NA | + | NA | NA |
| Goessinger et al, 2024 <sup>29</sup> | Prospective cohort study | Predictive | 205 | Melanoma | Combined | NA | + | NA | - |
| Hildebrand et al, 2023 <sup>30</sup> | Qualitative | Predictive | 15 | Metastatic cancer | Combined | +, - | + | NA | NA |
| Jutzi et al, 2020 <sup>16</sup> | Cross-sectional study | Predictive | 154 | Melanoma | Combined | +, - | NA | NA | NA |
| Kenig et al, 2024 <sup>41</sup> | Cross-sectional study | Predictive | 20 | Breast Cancer | NA | NA | + | NA | NA |
| Klotz et al, 2024 <sup>43</sup> | Cross-sectional study | Chatbot | 17 | Pancreatic Cancer | NA | NA | NA | + | NA |
| Lee et al, 2020 <sup>31</sup> | Descriptive | Predictive | 285 | Breast, Colorectal, Gastric, Gynecological, Liver, Lung, Thyroid | NA | NA | NA | + | NA |
| Leung et al, 2022 <sup>32</sup> | Pilot Study | Predictive | 48 | Breast, Gynecological, Colorectal, Head and Neck | NA | NA | NA | + | NA |
| Lyso et al, 2024 <sup>33</sup> | Qualitative | AI-NS | 30 | Prostate cancer | Combined | +, - | NA | NA | + |
| Manolitsis et al, 2023 <sup>34</sup> | Cross-sectional study | AI-NS | NR | Prostate cancer | NA | NA | + | NA | NA |
| McCradden et al, 2020 <sup>35</sup> | Qualitative | AI-NS | 18 | Meningioma | NA | - | U | NA | NA |
| Nally et al, 2024 <sup>42</sup> | Mixed methods study | AI-NS | 28 | Colorectal | Combined | +, - | - | NA | + |
| Nelson et al, 2020 <sup>17</sup> | Qualitative | AI-NS | 32 | Melanoma, Non-melanoma skin cancer | Combined | +, - | NA | NA | NA |
| Rodler et al, 2023 <sup>36</sup> | Cross-sectional study | Predictive | 257 | Prostate cancer | Combined | NA | + | NA | NA |
| Safran et al, 2024 <sup>37</sup> | Prospective study | Predictive, Chatbot | 166 | Breast, Colorectal | NA | NA | NA | + | NA |
| Temple et al, 2023 <sup>38</sup> | Cross-sectional study | Predictive | 95 | NR | NA | - | - | NA | NA |
| van Bussel et al, 2022 <sup>39</sup> | Mixed methods study | Chatbot | 135 | Breast, Colorectal, Gynecological, NR | NA | + | U | NA | NA |
| Yang et al, 2019 <sup>18</sup> | Cross-sectional study | Predictive | 402 | Lung, Breast, Pharyngeal, Lymphoma, others <sup>d</sup> | Combined | +, - | NA | NA | NA |

Abbreviations: AI, artificial intelligence; AI-NS, artificial intelligence - non specific; NA, not applicable; NR, not reported.
<sup>a</sup>Patient population with cancer in the included study only. For more detailed information on participant size and characteristics please see eTable in the Supplement.
<sup>b</sup>Sources were identified as either having a patient attitude that prefers: AI, physician, or a combined AI plus physician model.
<sup>c</sup>Predominant attitudes in each paper are indicated by Positive (+), Negative (-), or Undefined (U) to describe patients' feelings in respect to the given attitude.
<sup>d</sup>Top four types of cancer included in the study have been included in this list. For more detailed information please see eTable in the Supplement.

## Results

### General Characteristics of Included Studies

The 19 included studies, published between 2019 and 2025, represented 2,114 patients from 15 countries, with the majority originating from Germany, China, and South Korea.^16–18,28–43^ The majority of studies were published after 2022. Cancer types most frequently examined included prostate, melanoma, breast, colorectal, and lung cancers and are detailed in Table 2. Most studies employed quantitative methods (n=9), primarily using questionnaires or surveys, while qualitative studies (n=5) relied on interviews. The remaining mixed methods studies used a combination of surveys with both quantitative and qualitative answers in addition to interviews.

**Table 2.** Studies Included in the Scoping Review by Cancer Type and Patient Count

| Cancer Type | No. of Studies <sup>a</sup> | No. of Patients <sup>b</sup> |
| --- | --- | --- |
| Breast | 6 | 263 |
| Colorectal | 6 | 251 |
| Esophageal | 1 | 26 |
| Gastric | 2 | 25 |
| Gynecological | 4 | 60 |
| Liver | 2 | 39 |
| Lung | 2 | 135 |
| Lymphoma | 1 | 36 |
| Melanoma | 3 | 375 |
| Meningioma | 1 | 18 |
| Pancreatic | 1 | 17 |
| Pharyngeal | 1 | 40 |
| Prostate | 4 | 323 |
| Soft tissue sarcoma | 1 | 25 |
| Thyroid | 1 | 11 |
| Other | 5 | 63 |
| Unknown | 5 | 421 |

<sup>a</sup>Studies that include populations with multiple cancer types are listed under each cancer type for which they report findings.
<sup>b</sup>Includes data from one study<sup>18</sup> that included multiple tumor types for some patients.

## Review findings

### Attitudes

Patient attitudes were categorized into six key themes: preferences for AI-driven care, concerns about AI, optimism, trust, satisfaction, and fear and are summarized in Table 1.

### Preference for AI-Driven, Physician-Led or Combined Care

Nine studies explored patient preferences regarding care from AI only, a physician only, or combined AI-physicians using AI care models.^16–18,29,30,33,36,40,42^ The majority of patients preferred a combined model, in which physicians use AI as a decision-support tool rather than AI acting autonomously.^16–18,29,30,33,36,40,42^ One study reported that 83% of patients preferred AI-assisted screening over physician-only or AI-only screening.^29^ Patients valued AI’s ability to process large datasets, believing that its integration with physician expertise could improve clinical decision-making.^17,33,42^ However, a subset of patients expressed willingness to trust AI alone, particularly when it demonstrated superior diagnostic accuracy.^16,18,29,30,36,40^

### Concerns for AI Use in Care

Eight studies identified recurring concerns regarding AI integration.^16–18,30,33,35,38,42^ Patients frequently questioned the impact of AI on quality of care, with concerns that AI could lead to shorter appointment times, reduced patient-physician communication, and fewer opportunities for questions.^16–18,30,33,38,42^ Many also feared that AI could weaken the physician-patient relationship, contributing to depersonalized interactions.^16–18,33,38^ Concerns about AI’s susceptibility to false positives and false negatives were also raised, particularly regarding its lack of accountability in clinical decisions.^16–18,33^ Similarly, concerns about the collection and storage of patient data emerged, with patients emphasizing the need for explicit consent before data use and assurances that their information would be de-identified and protected from commercialization.^16,17,30,35^

### Optimism for AI Use in Care

Despite these concerns, seven studies highlighted optimism regarding AI’s role in their care.^16–18,30,33,39,42^ Many patients recognized AI’s potential to improve diagnostic accuracy, particularly in identifying subtle patterns that might be overlooked by physicians.^16–18,30,33,42^ AI was also seen as enhancing early detection and treatment efficiency, potentially reducing the need for invasive procedures and leading to less intensive treatment strategies.^16,33^ Additionally, AI was perceived as a tool that could improve the overall efficiency of their care and the healthcare system by reducing wait times, alleviating healthcare burdens, and lowering costs.^16–18,33^ Some patients believed that AI could also help reduce physician bias and ensure more consistent, data-driven treatment recommendations.^16,18,30,42^

### Trust

Trust in AI varied across studies.^29,30,34–36,38–42^ While some studies found high levels of trust, this was primarily dependent on whether AI was integrated with physician oversight.^29,34,35,41,42^ Multiple studies found that trust in AI was often rooted in physician endorsement rather than the AI itself, suggesting that patient trust in AI is closely tied to physician guidance.^30,36,42^ Education level played a role in AI trust; two studies found that patients with post-secondary education were more likely to participate in AI research.^34,40^ Education level was also correlated to previous exposure and understanding of AI technologies, which was another factor that positively influenced trust in AI.^18,40^ Conversely, another study found that older patients, and those with lower education levels were more likely to trust AI assessments.^29^

### Satisfaction

Five studies examined patient satisfaction with AI-assisted care.^28,31,32,37,43^ Patients reported greater satisfaction when AI improved diagnostic accuracy and increased healthcare efficiency.^31^ A study comparing an AI chatbot to specialists found that patients rated the AI’s responses slightly higher in empathy, comprehensibility, and content quality.^43^ One patient who used AI-driven emotional support tools, such as AI chatbots, reported reduced distress and anxiety following their cancer diagnosis.^28^ However, another study found that less than half of the participants in their study were satisfied with care received by AI, emphasizing the importance of integrating patients’ experiences into the development of AI-technologies.^37^

### Fear

Three studies explored fear as a patient response to AI integration.^29,33,42^ Some patients feared that AI could replace human interactions and limit their autonomy in decision-making.^33,42^ However, AI-driven diagnoses were better received when physicians confirmed the AI’s findings, underscoring the importance of physician oversight in maintaining patient trust.^29^

### Research User Engagement

As recommended by JBI and outlined in our protocol, we reviewed initial findings of this scoping review with research users at a BC Cancer Summit workshop.^24,26^ The workshop was attended by over 60 research users including clinicians, decision makers, researchers, and patients. Participants provided informal feedback, which broadly aligned with our findings, with many identifying more with the optimistic aspects of AI in cancer care. This engagement was intended to validate the relevance of our results and inform future research directions.

## Discussion

As AI adoption in cancer care accelerates, understanding patient perspectives is critical for ensuring its effective and ethical implementation. This scoping review provides the first synthesis of patient attitudes toward AI across all cancer types, revealing both areas of support and challenges in its integration in cancer care. While patients with cancer generally support AI when it is physician-guided, concerns about depersonalization, treatment bias, and data security highlight the need for careful implementation. This review identifies unique emotional and psychological concerns for patients with cancer, in contrast to the existing literature, which focuses more on general patients’ and the publics’ perspectives of AI, particularly related to its impact on diagnostic accuracy, efficiency, and cost-saving in healthcare.^15^

This review highlights a gap in understanding how AI is perceived across different stages of cancer, specifically among patients with metastatic disease. Despite AI’s potential to assist in prognosis and treatment planning, only 3 of the 19 included studies reported data on cancer staging, and patients with metastatic cancer represented less than 5% of the total study population.^30,31,36^ This underrepresentation limits insights into how AI is received by those with chronic and/or advanced cancer, and those receiving end-of-life care.^44,45^ Patients with metastatic disease may have distinct concerns regarding AI, including fears that AI-driven prognostic models could recommend less aggressive treatment options if predicted prognosis is poor, rather than prioritizing patient preferences.^12,13,30,46^ Future research should explore how AI can better support decision-making for patients with metastatic cancer, particularly in treatment planning, physician-patient communication, and prognostic discussions.

This review highlights a geographic imbalance in studies examining patient attitudes toward AI in cancer care. Most research has been conducted in Europe (n=1288)^16,29,33,34,36–43^ and Asia (n=688)^18,28,31^, with North American patient populations notably underrepresented (n=138)^17,30,32,35^. Studies from Asia have reported more optimistic attitudes toward AI, with some patients expressing trust in AI-assisted care without concerns.^18,31^ In contrast, studies from North America and Europe revealed more skepticism, with patients citing concerns related to over-reliance on AI^16,30,38^, treatment bias^30,33^, and data security^16,35^. These findings align with prior literature suggesting that trust and AI adoption are generally higher in Asia compared to Europe and North America.^39,47,48^ Future research should aim to incorporate more geographically diverse populations to ensure that AI implementation strategies reflect the values, concerns, and trust levels of patients across different healthcare settings.

The influence of educational background on patient attitudes toward AI in cancer care was underrepresented within the studies included in our review. While several studies explored the role of age and education on attitudes towards AI, a bias toward highly educated populations in these studies limited the generalizability of the findings.^16–18,29,30,34,35,40^ AI technologies have been shown to produce biased and inequitable outcomes across different patient backgrounds, and this underrepresentation in our review highlights a notable gap in research.^49,50^ It is essential that future research and AI implementation in cancer care incorporates the perspectives of diverse educational and socioeconomic groups to ensure that AI-driven care benefits all patients equitably.

Young adult patients with cancer were underrepresented in the studies included in this review. It is critical to understand the attitudes of this population as they have differences in disease biology, distribution, and survivorship compared to older patients with cancer.^51^ Greater exposure to AI technologies among younger patients may foster increased trust in its applications^18,34^, but some evidence suggests that a deeper understanding of AI’s complexity may also contribute to skepticism towards its diagnostic accuracy.^29,52^ Conversely, data suggesting that older patients were more likely to trust AI diagnoses raises concerns that misconceptions of AI technologies may result in misplaced trust and lead to uninformed or misguided decision-making around AI involvement in their cancer care.^29^ These generational differences in AI familiarity and usage will shape future patient attitudes as AI becomes more integrated into clinical practice. Further research is needed to ensure that perspectives of both younger and older populations are adequately represented in studies on AI implementation in cancer care.^51^

The successful integration of AI into cancer care depends on aligning these technologies with patient expectations and addressing concerns about trust, depersonalization, and ethical considerations. For AI researchers and developers, patients consistently express a preference for AI as a decision-support tool rather than an autonomous decision-maker.^16–18,29,30,33,36^ AI researchers and developers should prioritize human-AI collaboration models, ensuring that AI augments, rather than replaces, physician expertise. To improve the trust and usability of AI tools, researchers and developers should co-develop these tools alongside patients with cancer. Additionally, given the limited research on AI attitudes across different cancer stages, particularly metastatic cancer, future AI developments should consider how disease progression and prognosis influences treatment decisions. Developers must also recognize that AI acceptance varies globally, highlighting the need for regionally tailored AI implementation strategies that reflect diverse patient perspectives. Future AI development should incorporate the perspectives of young adult patients and individuals with diverse educational and socioeconomic backgrounds to promote more inclusive and patient-centered implementation.

For clinicians, clear communication and transparency about AI’s role in care are essential for building patient trust.^53^ Clinicians should proactively discuss how AI is being used, emphasizing that it supports rather than replaces their expertise. Preserving a strong physician-patient relationship is essential, as patients remain concerned that AI could depersonalize their care.^16–18,33,38^ Given that patient trust in AI is often linked to physician endorsement, clinicians can facilitate patient acceptance by integrating AI in a way that strengthens the human connection in care. Clinicians also play a role in the ethical implementation of AI by selecting tools that align with patient values and addressing concerns identified in this review, including depersonalization, bias, and data security. Lastly, clinicians can provide feedback to AI developers, ensuring that AI models are refined to better align with patient priorities and improve trust in AI-assisted care.

## Strengths and Limitations

This scoping review makes several novel contributions to the existing literature by being the first to comprehensively examine the attitudes of patients with cancer toward AI across all cancer types and treatment settings. While previous research has focused on AI acceptance in narrow contexts such as melanoma and breast cancer screening, this review provides a broader, more comprehensive synthesis of patient perspectives.^15–17^ Another strength of this review is its robust methodology, adhering to PRISMA-ScR guidelines, systematically identifying knowledge gaps, and synthesizing findings across diverse populations.^24^ Additionally, this review provides a patient-centered focus, offering insights that can inform AI development and guide clinical implementation.

Several limitations should be considered. As a scoping review, this study did not assess the quality of included articles, which is standard practice given that scoping reviews aim to map existing literature rather than evaluate methodological rigor. As a result, some of the included studies may vary in methodological quality. Also, the relatively small number of studies (n=19) included in the review may restrict the generalizability of our findings. This low number of studies is expected given the emerging nature of AI in cancer care. While this highlights a gap in the current literature, it also suggests that there is opportunity for further research to build on the findings presented in this review. Other limitations include the exclusion of non-English studies, secondary research, and grey literature. While secondary research could have offered relevant insights, we excluded it to avoid duplication and ensure that the review focused solely on primary literature. The exclusion of grey literature was necessary to maintain the review’s feasibility and manageability. Despite these limitations, the insights provided by this review offer a foundation for future research and will help guide the patient-centered implementation of AI into cancer care.

## Conclusions

This scoping review provides the first synthesis of patient attitudes toward AI across all cancer types, identifying both areas of support and concerns that impact its acceptance in clinical practice. While patients generally support AI when it complements physician decision-making, concerns about depersonalization, treatment bias, and data security highlight the need for careful implementation. Clinicians can use these findings to integrate AI in cancer care in ways that align with patient priorities, maintain human connection, and enhance trust, while researchers should address gaps in understanding AI perceptions among patients with advanced cancer and young adults.

## Supporting information

PRISMA-ScR

Flow Diagram

## Acknowledgements

The authors are grateful to the University of British Columbia librarian, Jane Jun, for her contributions to the search strategy.

The authors acknowledge the use of ChatGPT (GPT-4o, OpenAI) for language copyediting to enhance the fluency of the manuscript on February 1, 2025. All ideas, concepts, and original content were independently developed by the authors, and ChatGPT had no role in shaping the intellectual content. The authors take full responsibility for the accuracy and integrity of the manuscript.

## Supporting information

Supplementary Online Content - Search Terms and Summary of Included Studies PRISMA-ScR Checklist

## Authors Contributions

Concept and design: Daniel Hilbers, John-Jose Nunez, Navid Nekain

Acquisition, analysis or interpretation of data: Daniel Hilbers, Navid Nekain

Drafting of the manuscript: Daniel Hilbers, Navid Nekain

Critical revision of the manuscript for important intellectual content: Daniel Hilbers, Navid Nekain, John-Jose Nunez, Alan T. Bates

Administrative, technical, or material support: Daniel Hilbers

Supervision: John-Jose Nunez, Alan T. Bates

## Data Availability Statement

All relevant data from this study will be made available upon study completion.

