## Supplementary material for "Patient Attitudes Toward Artificial Intelligence in Cancer Care: A Scoping Review": Flow Diagram

**Identification of studies via databases and registers**

Records removed *before screening*:

Duplicate records removed by Covidence (n = 401)

Records from databases	(n = 1641):

MEDLINE (n = 957)

Embase (n = 358)

PsycINFO (n = 88)

CINAHL (n = 238)

**Identification**

Records screened

(n = 1240)

Records excluded

(n = 1167)

Reports sought for retrieval

(n = 73)

Reports not retrieved

(n = 0)

**Screening**

Reports excluded (n = 54):

Not primary research (n = 27)

Not patients with cancer (n = 9)

No patients’ attitudes (n = 7)

Mixed population: minority patients with cancer (n = 3)

Not available through public means or university library (n = 3)

No artificial intelligence (n = 2)

Duplicate (n = 2)

Not in English (n = 1)

Reports assessed for eligibility

(n = 73)

Studies included in review

(n = 19)

**Included**
