## Supplemental 1 for "Patient Attitudes Toward Artificial Intelligence in Cancer Care: A Scoping Review"

**Supplementary Online Content**

**eMethods.** Search Terms

**eTable.** Summary of Studies Included

**eMethods.** Search Terms

| **MEDLINE (OVID)** | |
| --- | --- |
| **Cancer** | (cancer* or neoplas* or malignan* or tumor*).tw,kw,kf. |
|  | OR |
|  | exp Neoplasms/ |
| AND | |
| **Artificial Intelligence** | (AI OR "artificial intelligence" or "computational intelligence" or "computer reasoning" or "computer vision systems" or ("knowledge Acquisition" adj2 computer*) or ("knowledge representation" adj2 computer*) or "Machine Intelligence" or "machine learning" or "natural language processing" or "computer neural network*" or "computational neural network*" or "connectionist model*" or "models neural network*" or "neural networks*" or perceptron* or "deep learning" or "hierarchical learning" or "data mining" or "text mining").tw,kw,kf. |
|  | OR |
|  | exp "Artificial Intelligence"/ or exp "Data Mining"/ |
| AND | |
| **Patients’ Perspective** | (patient* adj4 ("critical thinking" OR perception* OR perspective* OR emotion* OR feeling* OR regret* OR attitude* OR opinion* OR sentiment* OR "mental process*" OR trust OR distrust OR belief* OR experience*)).tw,kw,kf. |
|  | OR |
|  | Attitude/ or exp Emotions/ |
| **EMBASE (OVID)** | |
| **Cancer** | (cancer* or neoplas* or malignan* or tumor*).ab,kf,ti. |
|  | OR |
|  | exp Neoplasms/ |
| AND | |
| **Artificial Intelligence** | (AI OR "artificial intelligence" or "computational intelligence" or "computer reasoning" or "computer vision systems" or ("knowledge Acquisition" adj2 computer*) or ("knowledge representation" adj2 computer*) or "Machine Intelligence" or "machine learning" or "natural language processing" or computer neural network* or computational neural network* or connectionist model* or models neural network* or neural networks* or perceptron* or "deep learning" or "hierarchical learning" or "data mining" or "text mining").ab,kf,ti. |
|  | OR |
|  | exp "Artificial Intelligence"/ or exp "Data Mining"/ |
| AND | |
| **Patients’ Perspective** | (patient* adj4 ("critical thinking" OR perception* OR perspective* OR emotion* OR feeling* OR regret* OR attitude* OR opinion* OR sentiment* OR "mental process*" OR trust OR distrust OR belief* OR experience*)).ab,kf,ti. |
|  | OR |
|  | Attitude/ or exp Emotion/ |
| **PsychInfo (EBSCO)** | |
| **Cancer** | cancer* or neoplas* or malignan* or tumor* |
|  | OR |
|  | DE "Neoplasms" OR DE "Benign Neoplasms" OR DE "Breast Neoplasms" OR DE "Childhood Neoplasms" OR DE "Digestive System Neoplasms" OR DE "Endocrine Neoplasms" OR DE "Leukemias" OR DE "Lung Neoplasms" OR DE "Metastasis" OR DE "Nervous System Neoplasms" OR DE "Skin Neoplasms" OR DE "Terminal Cancer" |
| AND | |
| **Artificial Intelligence** | "AI" or "artificial intelligence" or "computational intelligence" or "computer reasoning" or "computer vision systems" or ("knowledge Acquisition" N2 computer*) or ("knowledge representation" N2 computer*) or "machine intelligence" or "machine learning" or "natural language processing" or "computer neural network*" or "computational neural network*" or "connectionist model*" or "models neural network*" or "neural networks*" or perceptron* or "deep learning" or "hierarchical learning" or "data mining" or "text mining" |
|  | OR |
|  | DE "Data Mining" OR DE "Text Analysis" OR DE "Artificial Intelligence" OR DE "Affective Computing" OR DE "Artificial Intelligence Ethics" OR DE "Cognitive Computing" OR DE "Computer Assisted Diagnosis" OR DE "Computer Linguistics" OR DE "Computer Vision" OR DE "Expert Systems" OR DE "Fuzzy Logic" OR DE "Heuristics" OR DE "Intelligent Agents" OR DE "Knowledge Representation" OR DE "Machine Learning" OR DE "Robotics" |
| AND | |
| **Patients’ Perspective** | patient* N4 ("critical thinking" or perception* or perspective* or emotion* or feeling* or regret* or attitude* or opinion* or sentiment* or "mental process*" or trust or distrust or belief* or experience*) |
|  | OR |
|  | DE "Attitudes" OR DE "Emotions" OR DE "Affective Valence" OR DE "Emotional Content" OR DE "Emotional Health" OR DE "Emotional Intelligence" OR DE "Emotional Processing" OR DE "Emotional Regulation" OR DE "Emotional Responses" OR DE "Emotional States" OR DE "Emotional Style" OR DE "Emotional Support" OR DE "Expressed Emotion" |
| **CINAHL (EBSCO)** | |
| **Cancer** | cancer* or neoplas* or malignan* or tumor* |
|  | OR |
|  | MH "Neoplasms+" |
| AND | |
| **Artificial Intelligence** | "AI" or "artificial intelligence" or "computational intelligence" or "computer reasoning" or "computer vision systems" or ("knowledge Acquisition" N2 computer*) or ("knowledge representation" N2 computer*) or "Machine Intelligence" or "machine learning" or "natural language processing" or "computer neural network*" or "computational neural network*" or "connectionist model*" or "models neural network*" or "neural networks*" or perceptron* or "deep learning" or "hierarchical learning" or "data mining" or "text mining" |
|  | OR |
|  | MH "Data Mining+" OR MH "Artificial Intelligence+" |
| **Patients’ Perspective** | patient* N4 ("critical thinking" or perception* or perspective* or emotion* or feeling* or regret* or attitude* or opinion* or sentiment* or "mental process*" or trust or distrust or belief* or experience*) |
|  | OR |
|  | MH "Attitude" OR MH "Emotions+" |

**eTable.** Summary of Studies Included

| **Citation** | **Journal** | **Journal Impact Factor** | **Study Population** | **Cancer Type(s)** | **Methodology** | **Key Findings** |
| --- | --- | --- | --- | --- | --- | --- |
| Au SCL. Patient with cancer who found support and care from ChatGPT. Cancer Res Stat Treat. 2023 Jun;6(2):305 | Cancer Research, Statistics, and Treatment | 0.67 | One patient with pre-existing cancer | Choroidal metastasis | A patient with cancer used ChatGPT to help cope with emotional stress after being diagnosed with cancer. | The patient felt lonely and afraid and looked to ChatGPT for comfort, counselling and guidance. ChatGPT provided him with support resources and left the patient feeling grateful. The patient felt comforted and supported after utilizing ChatGPT. |
| Fransen SJ, Kwee TC, Rouw D, Roest C, van Lohuizen QY, Simonis FFJ, et al. Patient perspectives on the use of artificial intelligence in prostate cancer diagnosis on MRI. Eur Radiol. 2025 Feb 1;35(2):769–75. | European Radiology | 4.7 | 212 patients undergoing MRI for prostate cancer (PCa) diagnosis or staging^a^ | Prostate Cancer | Patients received questionnaires on hypothetical AI assessment of their MRI scans. Opinions were assessed using a 5-point Likert-type agree-disagree scale. | 91% of patients wanted a second opinion from a radiologist after an AI MRI analysis of their scan. Trust in AI depended on education level and AI performance, if AI were to outperform radiologists, 52% would trust its results, while 43% were indecisive. Patients believed that the hospital (76%) and radiologist (70%) should take accountability for misdiagnosis. |
| Goessinger EV, Niederfeilner JC, Cerminara S, Maul JT, Kostner L, Kunz M, et al. Patient and dermatologists’ perspectives on augmented intelligence for melanoma screening: A prospective study. J Eur Acad Dermatol Venereol [Internet]. [cited 2024 Jun 8];n/a(n/a). Available from: https://onlinelibrary.wiley.com/doi/abs/10.1111/jdv.19905 | Journal of the European Academy of Dermatology and Venereology | 8.5 | 205 patients undergoing cancer screening and 8 dermatologists | Melanoma | Patients enrolled within an ongoing trial were screened for skin cancer by a dermatologist, followed by an independently operated AI 3D/2D total-body photography (TBP). The dermatologist provided a reassessment on all lesions knowing AI risks scores, which is referred to as an Augmented Intelligence (AuI). Patients were surveyed on their experiences and feelings following human, AI, and AuI examinations. | 95.5% of patients believed that AI could improve diagnostic performance. 5-fold increase in patient preference for human examination over AI alone. 42-fold increase in preference for synergistic application; 83.4% of patients preferred the AuI-based screening. Patients’ feelings of safety with AuI were significantly higher than AI or dermatologist screening alone.  The highest reduction in fear of developing skin cancer in patients was by the dermatologist’s exam (84.6%) followed by AI (2D-TBP=78.3%, 3D-TBP=77.9%). Patients’ trust in examinations by 2D/3D TBP was 92.9/92.3%, compared to 100% for the dermatologist’s examination. |
| Hildebrand RD, Chang DT, Ewongwoo AN, Ramchandran KJ, Gensheimer MF. Study of Patient and Physician Attitudes Toward Automated Prognostic Models for Patients With Metastatic Cancer. JCO Clin Cancer Inform. 2023 Jul;(7):e2300023 | JCO Clinical Cancer Informatics | 3.3 | 15 patients, 10 physicians, and 5 caregivers | Metastatic cancer | Semi structured interviews where participants were shown an anonymized prognosis and survival report for a patient with cancer from a Machine Learning survival model. Transcripts were coded, grouped thematically and analyzed using a constant comparative method. | All patients who completed the interview (n=14) wanted the option to have the ML model used in their care. Patients were interested by the wide number of factors the model considers and its comprehensiveness compared to physicians who are constrained by time demands. Patients trusted the model because it comes from physicians. Patient concerns included physician over-reliance on the model and bias to avoid treatment in the event of a shorter prognosis. |
| Jutzi TB, Krieghoff-Henning EI, Holland-Letz T, Utikal JS, Hauschild A, Schadendorf D, et al. Artificial Intelligence in Skin Cancer Diagnostics: The Patients’ Perspective. Front Med. 2020 Jun 2;7:233 | Frontiers in Medicine | 3.1 | 154 patients with previous diagnosis of melanoma and 143 patients with no melanoma history | Melanoma | Online survey evaluating patients’ expectations and concerns towards AI in general as well as attitudes toward other application scenarios. | 94% of respondents supported the use of AI in medical approaches. 91% believed AI should be used to support physicians to make more reliable skin cancer diagnostics. Top concerns around AI use were data protection, impersonality, susceptibility to errors, physician over reliance and loss of physician-patient relationships. |
| Kenig N, Muntaner Vives A, Monton Echeverria J. Is My Doctor Human? Acceptance of AI among Patients with Breast Cancer. Plast Reconstr Surg Glob Open. 2024 Oct 15;12(10):e6257. | Plastic and Reconstructive Surgery. Global Open | 1.5 | 20 patients who had undergone reconstruction for breast cancer | Breast Cancer | Patients part of a larger study involving AI and breast reconstruction evaluation were presented with a questionnaire about their attitudes towards AI. Questions were answered using a 1 (very low) through 10 (very high) scale. A free-text section was included in the questionnaire. | 65% scored their comfort with AI as very high. All patients rated their trust in AI as moderate to very high, with a majority (55%) scoring it as high (score of 7-8). Patients requested that physicians guarantee the accuracy of AI models, and did not wish for human doctors to be removed. |
| Klotz R, M. Pausch T, Kaiser J, et al. ChatGPT vs. surgeons on pancreatic cancer queries: accuracy & empathy evaluated by patients and experts. HPB. Published online December 2024. doi:10.1016/j.hpb.2024.11.012 | International Hepato-Pancreato-Biliary Association | 2.7 | 24 patient representatives and 25 surgeon specialists | Pancreatic Cancer | Frequently asked questions by pancreatic cancer patients were curated and independently solicited to ChatGPT, and two surgeons. Patients were asked to evaluate comprehensibility, context and empathy of responses using a 5 point Likert scale. | Patient responses showed that the AI Chatbot had the highest mean scores for content (4.05), lay comprehension (3.97), and empathy (3.24). 30% of the AI generated answers were selected as the “best answer” by patients. Surgeon responses held similar mean scores as AI but were selected as “best answer” more frequently (50%) than AI. |
| Lee K, Lee SH. Artificial Intelligence-Driven Oncology Clinical Decision Support System for Multidisciplinary Teams. Sensors. 2020 Aug 20;20(17):4693 | Sensors | 3.4 | 285 cancer patients receiving treatment | Breast, Colorectal, Gastric, Gynecological, Liver, Lung, Thyroid | Written questionnaires were given to patients receiving treatment from multi-disciplinary teams (MDTs) that either used the AI based Watson for Oncology (WfO) Clinical Decision Support System (CDSS) or did not. | Patients with MDTs that used AI were more satisfied by the treatment they received and had a more positive perception of the hospital after treatment (86.8% WfO vs 71.2% non-WfO). None of the participants reported negative attitudes. |
| Leung YW, Park B, Heo R, Adikari A, Chackochan S, Wong J, et al. Providing Care Beyond Therapy Sessions With a Natural Language Processing–Based Recommender System That Identifies Cancer Patients Who Experience Psychosocial Challenges and Provides Self-care Support: Pilot Study. JMIR Cancer. 2022 Jul 29;8(3):e35893 | Journal of Medical Internet Research Cancer | 5.8 | 48 online support group cancer patients | Breast, Gynecological, Colorectal, Head and Neck | Outputs from an AI based co-facilitator (AICF) used in online support groups were evaluated for precision and recall. Participants were surveyed on the usefulness of the AICF recommendation. | 76% of participants who received a resource from the AICF recommender system rated the resource they received as useful. |
| Lysø EH, Hesjedal MB, Skolbekken JA, Solbjør M. Men’s sociotechnical imaginaries of artificial intelligence for prostate cancer diagnostics – A focus group study. Soc Sci Med. 2024 Apr 1;347:116771 | Social Science and Medicine | 4.9 | 48 patients undergoing prostate cancer diagnosis | Prostate cancer | Focus groups discussing the topics of AI and prostate cancer diagnostics. Discussions focussed on general ideas about AI in prostate cancer diagnostics its potential. Reflexive thematic analysis was used to analyze the data obtained from transcripts. | Expectations for AI’s potential in prostate cancer diagnostics were simultaneously optimistic and pessimistic and were divided into either technologically centered or human centered. AI was seen as a tool that provides potential for more accurate diagnoses and personalized medicine. Trust of AI was not necessarily the technology, but rather the healthcare system that uses the technology. Patients were fearful that AI would degrade communicative situations and exclude them from the decision-making process. |
| Manolitsis I, Tzelves L, Feretzakis G, Kalles D, Verykios VS, Katsimperis S, et al. Acceptance of Artificial Intelligence in Supporting Cancer Patients. In: Mantas J, Gallos P, Zoulias E, Hasman A, Househ MS, Charalampidou M, et al., editors. Studies in Health Technology and Informatics [Internet]. IOS Press; 2023 [cited 2024 Nov 9]. Available from: https://ebooks.iospress.nl/doi/10.3233/SHTI230561 | Studies in Health Technology and Informatics | 0.68 | Prostate cancer patients undergoing radical prostatectomy  (number unknown) invited to participate in ASCAPE study | Prostate cancer | Patients undergoing radical prostatectomy, who were invited to an international study on AI for quality-of-life issues in prostate cancer survivors (ASCAPE), were asked to answer questions about baseline demographics, perception on the use of internet, smartphone applications, and AI in general. Participants included those who declined to participate in the ASCAPE study. | Almost all patients who chose to participate in the ASCAPE study agreed with the use of AI in medicine. Of those that declined participating in the study, about one third disagreed with the use of AI in medicine. Prostate cancer patients that accepted to participate in the ASCAPE study were younger, lived in urban centres and had completed higher education. |
| McCradden MD, Baba A, Saha A, Ahmad S, Boparai K, Fadaiefard P, et al. Ethical concerns around use of artificial intelligence in health care research from the perspective of patients with meningioma, caregivers and health care providers: a qualitative study. CMAJ Open. 2020 Feb 11;8(1):E90 | CMAJ Open | 12.9 | 18 cancer patients, 7 caregivers, 5 health care providers part of larger quality-of-life study | Meningioma | Qualitative interviews were conducted with participants based on vignettes of realistic but hypothetical AI-enabled research and ethical issues. | Participants expressed a mixture of concern and optimism towards the use of AI in healthcare research. Although there was broad support for the use of AI in medicine, concerns included those around privacy, consent, confidentiality, responsibility, accountability, harm and trust. |
| Nally DM, Kearns EC, Dalli J, Moynagh N, Hanley K, Neary P, et al. Patient public perspectives on digital colorectal cancer surgery (DALLAS). Eur J Surg Oncol. 2024 Oct 20;108705. | European Journal of Surgical Oncology | 3.5 | 28 cancer patients who had received treatment and surgery, 8 family members | Colorectal | Focus groups discussing the topics of research, data, industry, and AI in surgery were conducted across two separate events. Inductive thematic analysis with a reflexive approach was used to perform qualitative analysis. Pre and post surveys utilizing Likert scale and free text answers were used to further record patients’ perspectives. | Principal attitude towards AI was defined as “AI needs human input.” Subthemes of fear and misconceptions towards AI were identified, as well as a need for human control over final decisions when there is AI involvement. 80% of participants agreed that AI should be applied in surgical care, but felt the surgeon was more valuable than AI. Participants felt that AI should not make diagnoses (64%) or treat patients (70%) without human input. |
| Nelson CA, Pérez-Chada LM, Creadore A, Li SJ, Lo K, Manjaly P, et al. Patient Perspectives on the Use of Artificial Intelligence for Skin Cancer Screening: A Qualitative Study. JAMA Dermatol. 2020 May 1;156(5):501–12 | JAMA Dermatology | 11.5 | 48 dermatology patients | Melanoma, Non-melanoma skin cancer | Qualitative semi structured interviews exploring direct-to-patient and clinician decision-support AI tools. Grounded theory approach was used to develop the codebook, and interviews were performed until thematic saturation. | 94% of participants believed in the importance of a symbiotic relationship between AI and humans. 75% would recommend an AI tool to family or friends. Diagnostic speed (60%) and health care access (60%) were the two most common perceived benefits of AI for skin cancer screening. Both accuracy (69%) and lack of accuracy (85%) were identified as AI’s greatest strength and weakness. In the event of conflicting diagnosis 67% of patients would seek a biopsy, but 60% would still put more trust in a doctor. |
| Rodler S, Kopliku R, Ulrich D, Kaltenhauser A, Casuscelli J, Eismann L, et al. Patients’ Trust in Artificial Intelligence–based Decision-making for Localized Prostate Cancer: Results from a Prospective Trial. Eur Urol Focus. 2023 Nov;S2405456923002377 | European Urology Focus | 4.9 | 466 patients receiving diagnostic or therapeutic interventions for prostate cancer | Prostate cancer | Patients receiving interventions were given a questionnaire assessing their perspectives on AI implementation in clinical workflows during PC treatment. 5-point Likert scale was used to assess trust in AI. | 67.2% of patients would choose an AI-assisted physician for treatment over either physician or AI alone. Patients’ trust in AI was slightly positive (3.41 ± 1.19), with significant positive correlation with cumulative affinity for technology. When making a diagnosis, patients trusted an AI controlled by a physician (4.31 ± 0.88) the most. Patients trusted physicians’ to make the right diagnosis, integrate new research, provide individualized responses and explain information over that of an AI. In the case of disagreement between AI and physician, 70.8% would trust the physician over AI. |
| Šafran V, Lin S, Nateqi J, Martin AG, Smrke U, Ariöz U, et al. Multilingual Framework for Risk Assessment and Symptom Tracking (MRAST). Sensors. 2024 Feb 8;24(4):1101 | Sensors (Basel, Switzerland) | 3.4 | 166 breast and colorectal cancer survivors part of previous trial | Breast, Colorectal | Patients using the Multilingual Framework for Risk Assessment and Symptom Tracking (MRAST) framework’s mHealth app were given app-based questionnaires at three distinct time intervals to gain insights into their experience in the study. These included the mHealth app, video diaries, and chatbot questionnaires. Patients rated their experience on a scale of 1 (poor) to 10 (excellent). | Patients’ experiences with the mHealth app, chatbot questionnaires and diary recordings was positive for the duration of the study. Most participants experienced usability issues, with only 44% of patients rating the usability of the system as good or excellent by the end of the study. 75.3% of patients were willing to engage in health monitoring activities using digital wearable devices. |
| Temple S, Rowbottom C, Simpson J. Patient views on the implementation of artificial intelligence in radiotherapy. Radiography. 2023 May 1;29:S112–6 | Radiography | 2.5 | 130 cancer patients receiving radiotherapy | N/A | An existing questionnaire using 5-point Likert-type scales was adapted to assess patients’ views on implementation of AI in radiotherapy and distributed to patients after treatment completion. | Patients’ trust in AI for use in radiotherapy was moderately negative (Distrust: 3.24 ± 0.66). Patients had a strong desire to understand procedural knowledge in radiotherapy and to be able to talk to someone about it (4.39 ± 0.55). Personal interactions were similarly highly valued (4.36 ± 0.49). |
| van Bussel MJP, Odekerken–Schröder GJ, Ou C, Swart RR, Jacobs MJG. Analyzing the determinants to accept a virtual assistant and use cases among cancer patients: a mixed methods study. BMC Health Serv Res. 2022 Jul 9;22(1):890 | BMC Health Services Research | 2.7 | 8 former patients with cancer, 127 cancer treatment recipients across Europe and North America | N/A | Qualitative interviews with former cancer patients and health care workers around acceptance factors for a Virtual Assistant (VA) were conducted and analyzed to form a quantitative self-administered online survey. Patients awaiting or receiving treatment were invited to participate in the survey. | Performance expectancy, effort expectancy, social influence and trust were the leading factors that influenced a patient’s intention to use a VA. The interviewed patients found many aspects of the VA useful, including the ability to answer logistic, treatment procedure, side effect and scheduling questions, and could reduce anxiety before treatment. |
| Yang K, Zeng Z, Peng H, Jiang Y. Attitudes Of Chinese Cancer Patients Toward The Clinical Use Of Artificial Intelligence. Patient Prefer Adherence. 2019;13:1867–75 | Patient Preference and Adherence | 2.0 | 402 cancer patients | Pharyngeal, Lung, Breast, Esophageal, Gastric, Colorectal, Liver, Lymphoma, Cervical, Soft tissue sarcoma, Others^b^ | Questionnaire investigating patients’ attitudes towards AI in Medicine (AIM) was distributed to eligible cancer patients. | 90% believed in a diagnosis provided by an AI doctor, but 88.8% would prefer the diagnosis of a human doctor in the event of a conflicting diagnosis. Patients had similar views about therapeutic advice given by AIM. Post treatment, 88.3% would prefer to discuss their therapy or prognosis with a human doctor. The major disadvantage of AIM identified was a lack of capacity to deal with complicated disorders. AIM was seen as economical and convenient by most patients. 56.7% believed that AI would assist physicians as opposed to 30.3% that believed physicians would assist AI doctors. |

^a^Although all patients were being assessed for screening MRI, 36 reported a previous history of prostate cancer, and 170 for other cancers.

^b^Total count of tumor types (n=416) in this population exceeds actual population (n=402), as some patients reported multiple tumor types.
